# Familial risk of postpartum psychosis

**DOI:** 10.1101/2023.07.20.23292910

**Authors:** Adrianna P. Kępińska, Thalia K. Robakis, Keith Humphreys, Xiaoqin Liu, René S. Kahn, Trine Munk-Olsen, Veerle Bergink, Behrang Mahjani

## Abstract

**Objective:** Postpartum psychosis, a mood disorder triggered by childbirth, is one of the most severe psychiatric conditions, with high risks of suicide and infanticide if untreated. While it is evident that genetic factors play a crucial role in disorder risk, the exact extent of their importance is yet to be determined.

**Methods:** This cohort study consisted of 1,648,759 women from the Swedish nationwide registers, of whom 2,514 (0.15%) experienced postpartum psychosis within three months of their first-ever childbirth. We estimated the relative recurrence risk of postpartum psychosis for female full siblings and cousins as a measure of familial, genetic, and environmental risk.

**Results:** Relative recurrence risk of postpartum psychosis in full siblings was 10.69 (95% CI=6.60–16.26) when adjusted for year of and age at childbirth. Although cousins showed an elevated relative recurrence risk, these results did not reach statistical significance (1.78, 95% CI=0.70–3.62). Despite the higher familial risk of postpartum psychosis among full siblings, the absolute risk for women with an affected sibling is relatively low, estimated at 1.55% within the entire population.

**Conclusions:** The observed increased risk of postpartum psychosis in full siblings suggests both genetic and shared environmental influences. However, the lack of significant results in cousins hampers a definitive distinction between these factors. Furthermore, despite increased relative recurrence risk in siblings, their overall likelihood of developing postpartum psychosis remains low. Our study underscores the need for further research to better understand the intricate interplay of genetics and environment in the development of postpartum psychosis.

## 1. Introduction

Postpartum psychosis is one of the most severe psychiatric conditions, with increased risks of suicide and infanticide if untreated, and should thus be addressed as a medical emergency (1). Postpartum psychosis usually occurs in the three months after childbirth, with the most frequent onset within one month (2–4). The onset is rapid, and presentation is fluctuating, characterized by mood symptoms (mania and depression), psychotic symptoms, cognitive disorganization and confusion, severe anxiety, and sleep problems (1). Despite its significant impact, postpartum psychosis continues to pose a challenge in terms of diagnosis and treatment, as it is not acknowledged as a separate disorder in the Diagnostic and Statistical Manual of Mental Disorders (DSM)(1, 5).

Notably, the risk architecture of postpartum psychosis is insufficiently researched. As early as in one of the first systematic clinical monographs on women’s perinatal mental health, *Treatise on Insanity in Pregnant, Postpartum, and Lactating Women* (1858), Louis-Victor Marcé theorized that individuals might be predisposed to mental illness through “hereditary antecedents,” with pregnancy, delivery, or lactation as disorder triggers. However, while some research now indeed points to biological factors in postpartum psychosis, e.g., structural brain changes (6), neuroimmune dysregulation (7, 8), and changes in the hypothalamic-pituitary-adrenal axis (9), genetic findings remain largely absent. Earlier research includes family studies of postpartum psychosis, narrowly in patients with a history of bipolar disorders (10, 11). A significant limitation of these studies lies in the challenge of determining the causes of postpartum psychosis in families with a history of bipolar disorder. Specifically, it is difficult to ascertain whether the frequent occurrence of postpartum psychosis in these families results from shared genetic factors, common environmental influences, or a combination of both. Additionally, it is important to acknowledge that these studies inherently involve a highly selected sample of individuals who, by design, are at a higher risk of developing postpartum psychosis. More recent studies include candidate gene studies (e.g., (12)) and a single study applying the now widely used methodology of polygenic scoring in women with bipolar disorder and postpartum psychosis (13). Conducting research in this area remains challenging due to the rarity of postpartum psychosis, making the acquisition of sufficiently large samples a difficult task.

Here, we provide a population-based cohort study of relative recurrence risk of postpartum psychosis in female full siblings and cousins of women who had experienced postpartum psychosis following their first liveborn childbirth. We used data from Swedish national registers to measure familial risk of postpartum psychosis by estimating relative recurrence risk. Relative recurrence risk is commonly used in genetic and family-based studies to quantify the risk of a specific condition occurring in one individual within a family based on the presence of the same condition in another family member. The term “relative recurrence risk” emphasizes familial aggregation of the condition. Relative recurrence risk does not imply the actual recurrence of the condition within the same individual but rather the increased likelihood of its appearance in another individual within the family (recurrence in the family). Relative recurrence risk can be used to compare the risk of the condition among families to the risk observed in unrelated individuals, i.e., population prevalence.

In this study, we compared relative recurrence risk across different family relationship types. In light of previous findings that postpartum psychosis occurred more in women with bipolar disorder and a family history of postpartum psychosis in a first-degree relative, compared to women with bipolar disorder but no family history (10, 11), we hypothesized that relative recurrence risk for postpartum psychosis varies by degree of genetic relatedness and is higher in female full siblings than in cousins.

## 2. Methods

### 2.1 Study population

This cohort study used data from Swedish national registers. Swedish healthcare covers all residents, with their healthcare data systematically collected in these national registers. Information across registers is linked using individuals’ personal identity numbers.

We used data from the Medical Birth Register (MBR)(14) to select all women who had their first-ever liveborn childbirth between January 1, 1980, and October 31, 2017. We followed women from their own birth until December 31, 2017. The MBR was established in 1973 and contains birth and neonatal information on approximately 98% of all births in Sweden. The data quality of the MBR is high, owing to semi-automated data extractions from resident electronic health records and data quality controls for completeness and errors by the government agency National Board of Health and Welfare (Socialstyrelsen)(14).

It is important to note that our study utilizes registry data based on biological sex assigned at birth. We recognize that this designation may not correspond to the current gender identity of all individuals in the dataset. For the purposes of this study, the terms ‘female,’ ‘women,’ and ‘mother’ refer to individuals assigned female at birth, as recorded in the registry data.

To identify full sibling and cousin pairs, we extracted family information from the Multi-Generation Register (MGR)(15), focusing on female (i.e., same-sex) full sibling and cousin relationships. In this context, “cousins” refers to first degree cousins. MGR collects information on individuals born in the country since 1932, has its origins in personal records established in 1947. The MGR includes not only personal records but also details on the relationships of individuals to their biological and adoptive parents. It has a coverage of 97% of mothers and 95% of fathers of index individuals (16). The data quality of the MGR is also high, as data are regularly audited for errors and cross-checked with other Swedish registers by the government agency Statistics Sweden (15).

Ethical approval and waiver of informed consent (which is not required for the use of register data) were obtained from the Regional Ethical Review Board in Stockholm, Sweden.

### 2.2 Outcomes and Exposure Covariate

In this study, our primary outcome of interest was the diagnosis of postpartum psychosis. We obtained diagnostic information for psychiatric disorders from the Swedish National Patient Register (NPR), which includes both inpatient and outpatient care data.

The inpatient data are sourced from the National Inpatient Register (IPR; Swedish: slutenvårdsregistret), also known as the Hospital Discharge Register. Established in 1964 and achieving nationwide coverage in 1987, the IPR is a crucial component of the NPR and currently registers more than 99% of all somatic and psychiatric hospital discharges in Sweden.

In addition to the inpatient records from the IPR, our study also leveraged outpatient data reported to the Swedish National Patient Register (NPR) from the Outpatient Care Register (Swedish: öppenvårdsregistret). Since 2001, Swedish counties have been required to report hospital-based outpatient physician visits. While inpatient care coverage in the IPR is nearly complete, nearing 100%, outpatient care coverage is relatively lower, primarily due to incomplete data capture from private healthcare providers. (17). Nonetheless, the coverage for outpatient care provided by public caregivers is nearly as comprehensive as inpatient care. Since 2006, the proportion of missing primary outpatient diagnoses has been consistently decreasing, and in psychiatry, has been at less than 10% since 2013 (18). This indicates that, while the NPR likely captures the majority of psychiatric cases, some cases, potentially including those of postpartum psychosis, may still be missing.

Diagnoses within the NPR are made by clinical specialists and are recorded using the International Classification of Diseases (ICD) codes: ICD-8 from 1968 to 1986, ICD-9 from 1987 to 1996, and ICD-10 since January 1, 1997. Our study leveraged both primary and secondary diagnoses from the NPR, providing a comprehensive overview of psychiatric conditions affecting the study population.

#### Postpartum psychosis

Postpartum psychosis is often considered an umbrella term (19), lacking official diagnostic criteria in major classification systems like the Diagnostic and Statistical Manual of Mental Disorders (DSM). In the ICD system, “puerperal psychosis” is coded as ICD-8 294.4 and ICD-10 F53.1, categorizing “severe mental and behavioural disorders associated with the puerperium, not elsewhere classified.” These Not Otherwise Specified (NOS) codes are used when a diagnosis lacks specific details, allowing for a broad categorization of mental health conditions when the exact nature of the disorder is unclear or inadequately documented. Our analysis revealed no instances of the ICD-8 294.4 code being used and limited application of the ICD-10 F53.1 code for postpartum psychosis (190 patients), highlighting the necessity for clinicians to rely on comprehensive clinical judgment and assessment beyond the ICD categorization to accurately diagnose postpartum psychosis.

Thus, to determine postpartum psychosis cases, we used the diagnostic classification, which has been widely used in the literature: patients with mania and/or psychosis with onset postpartum (5). For the main analysis, we selected disorders with symptom onset present in women within 0-3 months following their first liveborn childbirth. We selected the period of up to three months to provide a broad coverage of the disorder onset period (2–4, 20, 21).

To cover all episodes with mania and psychosis with onset postpartum, we included the following diagnoses using ICD-8, 9, and 10 codes: “brief psychotic disorder; psychotic disorders not due to a substance or known physiological conditions; manic episode; bipolar disorder; major depressive disorder, single episode, severe with psychotic features; recurrent depressive disorder, current episode severe with psychotic symptoms features; and puerperal psychosis “(ICD codes are listed in Table S1 in the Supplement). Throughout this manuscript, diagnosis of postpartum psychosis refers to this combined approach of using multiple ICD codes, ensuring a comprehensive and consistent identification of cases in line with recognized psychiatric classifications.

#### Covariates

We included three covariates in the analyses. The first one was year of the first-ever childbirth (birthyear), to account for variation in dataset completeness, as historical family records may potentially be less complete in earlier years of the registers (for more details, see missing data section). The second covariate was the age of the mother at first-ever childbirth (measured in years), given that older age at delivery has been linked to an increased risk of hospital admissions for postpartum psychosis among Swedish first-time mothers (22). The third covariate was a history of any diagnosis of bipolar disorder before the first-ever childbirth.

### 2.3 Statistical analysis

To investigate the familial aggregation of postpartum psychosis, we focused on estimating the relative recurrence risk among female full siblings and cousins. This measure indicates the increased likelihood of developing postpartum psychosis for individuals with a family history of the condition compared to those without. We defined exposure as having a female sibling or cousin previously diagnosed with postpartum psychosis. By comparing the occurrence of postpartum psychosis in siblings/cousins (affected family members) to those without a familial history, we aim to elucidate the influence of familial presence of the condition on individual risk.

The relative recurrence risk can be calculated in different ways depending on the study design and available data. One approach involves comparing the prevalence of a condition in individuals with a family history to the prevalence in the general population. This approach relies on accurate population prevalence data, which may be limited or outdated. Furthermore, this approach does not account for other confounding factors. Another approach involves logistic regression, which estimates an odds ratio, indicating the relative increase or decrease in the odds of a condition occurring in individuals with a family history of a condition compared to those without. This approach does not rely on population prevalence and allows for an adjustment of potential confounding factors in the model. However, it provides a sample-specific estimate of relative recurrence risk and may not facilitate direct comparisons across different populations.

In this study, we used the latter approach, logistic regression, to calculate the relative recurrence risk of postpartum psychosis in full siblings/cousins. This risk represents the likelihood of individuals with a female sibling/cousin having a history of postpartum psychosis developing the condition, compared to individuals with a sibling/cousin without a history of postpartum psychosis. First, we extracted all pairs of full siblings (sibling1, sibling2). We performed logistic regression by using sibling1 disease status as the predictor and sibling2 disease status as the outcome (with and without adjustment for the sibling2 age at childbirth and history of bipolar disorder covariates). We then repeated these analyses for all pairs of cousins (cousin1, cousin2) with postpartum psychosis diagnosis. We performed the analyses for all individuals who received a diagnosis of postpartum psychosis (in either inpatient or outpatient care). We calculated a 95% profile likelihood confidence interval for each parameter.

We conducted all analyses with R version 4.0. To calculate relative recurrence risk, we used two functions from the R package *stats*: the *glm* function to fit logistic regressions and the *confint* function to obtain 95% confidence intervals of the exponents of the parameter estimates.

#### Missing data

We encountered two main challenges concerning data completeness in our study. First, since postpartum psychosis exclusively affects women, our analysis is inherently limited to families where women have female siblings or cousins eligible for inclusion.

Secondly, the completeness of historical records poses a challenge, particularly when identifying cousin relationships that require data spanning multiple generations. To estimate the number of missing cousins, we analyzed siblings’ and cousins’ birth years, spanning 1934-2002 and 1953-2001, respectively. We divided the data by decade to evaluate data completeness over time. The 1980-1990 period, showing the highest cousin-to-sibling ratios, was selected for its more complete records, likely due to improved record-keeping practices. Using the 1980-1990 ratio as a baseline, we calculated the expected number of cousins for each decade and compared these to actual counts. This comparison revealed an overall missingness of approximately 39% in our dataset.

#### Sensitivity analyses

We conducted two sensitivity analyses to test the robustness of the relative recurrence risk estimate. First, we used a subsample with inpatient diagnoses only to address the severity of postpartum psychosis. We defined an inpatient diagnosis of postpartum psychosis if any of the diagnoses mentioned above, recorded with the ICD codes, was given during a stay in an inpatient care unit within three months of the first-ever childbirth. Conversely, if diagnostic codes were assigned in outpatient care only, we classified the diagnosis as an outpatient diagnosis of postpartum psychosis. Importantly, since outpatient data have been consistently available since 2001, we limited our analysis to childbirths occurring after this year to ensure comparability. This approach allowed us to assess the recurrence risk in a dataset that uniformly reflects both inpatient and outpatient cases from the period when outpatient data was systematically recorded (Figure S1).

Additionally, in our sensitivity analysis that examines data from 2001 onwards, we focus on a critical period in our dataset: the children born in 2001, who typically have mothers born around 1984. This year marks a significant juncture in our assessment, aligning with the period we identify as having complete records. The choice of this timeframe allowed us to indirectly assess the impact of missing data on cousin relationships with a higher degree of confidence in the data completeness.

For the second sensitivity analysis, to ensure consistency in definitions across various versions of ICD codes, we limited the sample to individuals with ICD-10 diagnosis codes only and recalculated the relative recurrence risk estimates. Given that ICD-10 was adopted in 1997 (18), this decision inherently limits our cohort to childbirths that occurred after 1997.

## 3. Results

The cohort included all liveborn childbirths in Sweden between January 1, 1980, and October 31, 2017, and followed up to December 31, 2017. The cohort included 1,648,759 women, of whom 2,514 (0.15%) experienced postpartum psychosis within three months of their first-ever childbirth (Figure 1 and Tables 1-3).

**Figure 1.**
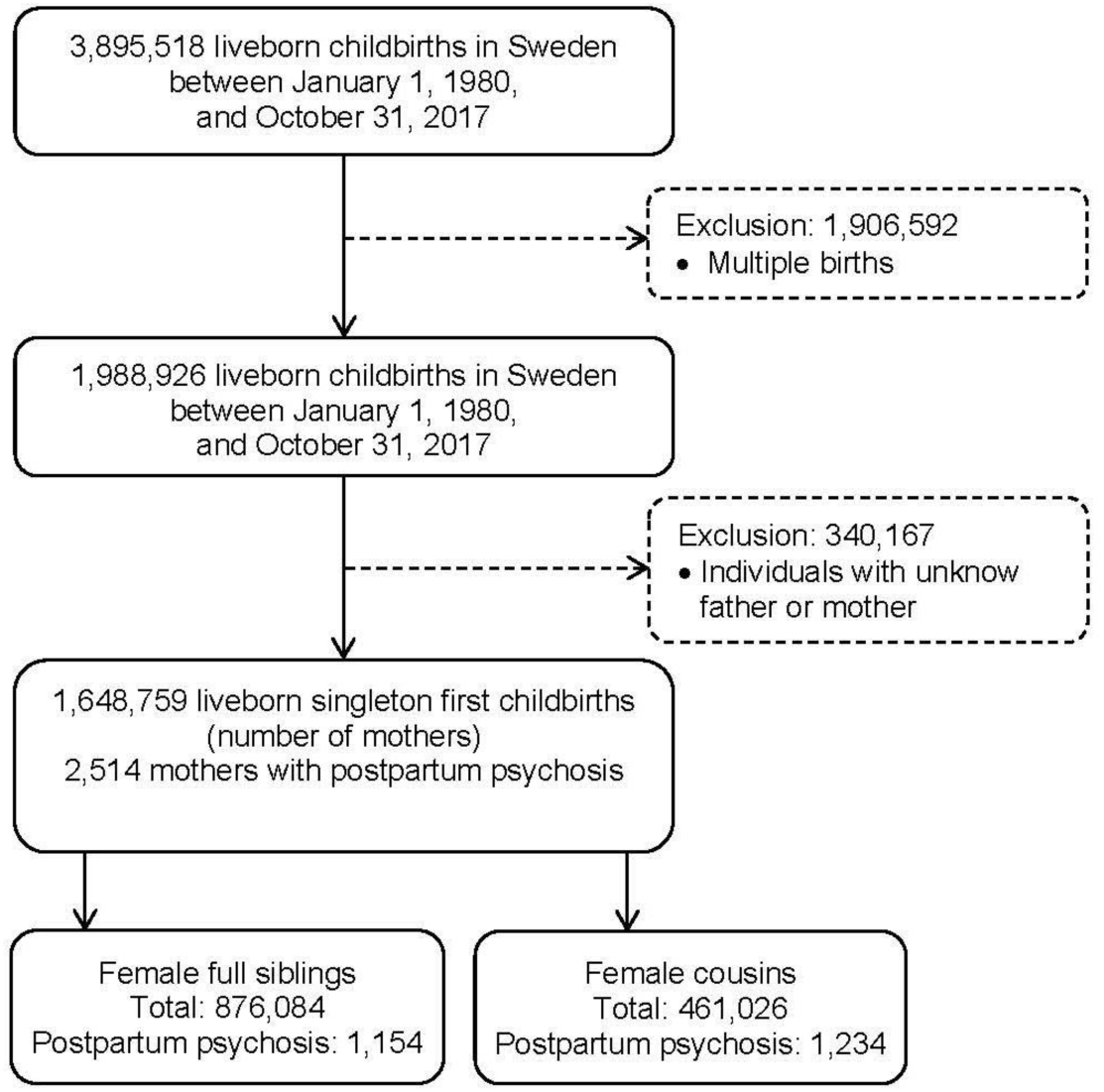
Flow chart of the study population selection.

**Table 1.** Sample characteristics of mothers with a first-ever childbirth between January 1, 1980, and October 31, 2017, and postpartum psychosis diagnosis based on ICD-8, 9 and 10.

| Characteristic | Full Sample<br>(N = 1,648,759) |  | Full Siblings<br>(N = 876,084) |  | Cousins<br>(N = 461,026) |  |
| --- | --- | --- | --- | --- | --- | --- |
|  | N | % | N | % | N | % |
| Postpartum psychosis diagnosis | 2,514 | 0.15 | 1,154 | 0.13 | 1,234 | 0.27 |
| History of bipolar disorder during the study period | 22,993 | 1.40 | 11,855 | 1.40 | 8,435 | 1.80 |
| History of bipolar disorder before childbirth <sup>a</sup> | 3,747 | 0.23 | 1,520 | 0.18 | 2,612 | 0.56 |
| History of bipolar disorder before childbirth among mothers with a postpartum psychosis diagnosis <sup>b</sup> | 1,234 | 49 | 511 | 44 | 832 | 67 |
| History of a mental disorder during the study period <sup>c</sup> | 179,149 | 10.90 | 91,128 | 10.40 | 55,144 | 12.00 |
| History of a mental disorder before childbirth <sup>c</sup> | 11,383 | 0.69 | 5,277 | 0.60 | 4,808 | 1.04 |
| History of a mental disorder before childbirth among mothers with a postpartum psychosis diagnosis <sup>b,c</sup> | 1,839 | 73 | 799 | 69 | 1,009 | 82 |
|  | Mean | SD | Mean | SD | Mean | SD |
| Age at childbirth (years) | 29.3 | 5.3 | 29.0 | 5.1 | 27.4 | 4.5 |

<sup>a</sup> Used as a covariate in the analysis.
<sup>b</sup> Percentage breakdown of the diagnoses is the percentage out of the total number of postpartum psychosis cases.
<sup>c</sup> Cases with history of a bipolar disorder diagnosis are included in the total of cases with history of a mental disorder diagnosis.
Note: Each individual can belong to both full sibling and cousin family relationships.

During the study period (from each mother’s own birth up to December 31, 2017), 179,149 individuals (10.9% of the cohort) had a diagnosis of any mental disorder, according to ICD codes (Table S2). Within the group diagnosed with postpartum psychosis, 1,234 (49%) had a history of bipolar disorder and 605 (24%) had other mental health diagnosis before their first childbirth, making a total of 1,839 (73%) with a pre-existing mental disorder diagnosis. The prevalence of bipolar disorder in our cohort was 1.4% (22,993 individuals). These findings underscore a very high risk of postpartum psychosis in patients with history of bipolar disorder. This highlights the importance of prevention of these severe postpartum episodes in pregnant individuals.

We identified a discernible upward trend in the prevalence of postpartum psychosis over time (Figure S2). This trend could inflate the estimates of relative recurrence risk. Additionally, the increasing prevalence over time poses a challenge in analyzing cousin relationships compared to full siblings. Tracing family trees back in time becomes increasingly challenging, as data from the early years of registers are often less complete than more recent patient data. As a result, cousin data are disproportionately influenced by the higher prevalence rates observed in these later periods compared to full siblings (Figure S1). Consequently, we anticipated observing higher rates of postpartum psychosis among cousins compared to siblings. To address these challenges and ensure our analysis accounts for these temporality and data completeness issues, we adjusted all our analyses for the birthyear of the first child.

In full siblings with postpartum psychosis following childbirth, compared to those with no diagnosis, relative recurrence risks were high and statistically significant (relative recurrence risk, adjusted for birthyear = 10.34, 95% CI=6.58–16.20, p<0.001) and substantially lower and not statistically significant in cousins (relative recurrence risk, adjusted for birthyear = 1.82, 95% CI=0.72–3.72, p=0.143). This pattern was similar for models adjusted for birthyear and age at childbirth (Tables 4 and S3). In the model adjusted for birthyear, age at childbirth, and siblings/cousins history of bipolar disorder before birth, the relative recurrence risk remained significant for full siblings but decreased relative to the model unadjusted for history of bipolar disorder before birth (adjusted relative recurrence risk = 6.88, 95% CI=3.50–12.71, p<0.001). Additionally, the coefficient for the history of bipolar disorder in this model was 0.73 which corresponds to odds ratio of 2.08 (95% CI=1.03–4.01, p=0.036), quantifying the increased risk of postpartum psychosis in a sibling based on the presence of bipolar disorder in the other sibling. In cousins, in the model adjusted for age at childbirth and cousin’s history of bipolar disorder, the relative recurrence risk of postpartum psychosis was not significant (with a confidence interval that does not exclude 1).

An analysis limited to the period after 2001, where data reliability and familial link completeness are enhanced, yielded a slightly decreased risk estimate for full siblings of 7.82 (95% CI=3.88–13.95, p<0.001), while the risk for cousins remained consistent at 1.79 (95% CI=0.71–3.66, p=0.141).

In full siblings, we observed that the relative recurrence risk for a potentially more severe form of postpartum psychosis, requiring inpatient admission (adjusted for age at childbirth), did not show a meaningful difference compared to the relative recurrence risk for postpartum psychosis encompassing both inpatient and outpatient cases, as indicated by the widely overlapping confidence intervals (Tables 2, 3, S3, and S4).

**Table 2.** Sample characteristics of mothers with a first-ever childbirth between January 1, 2001, and October 31, 2017, and postpartum psychosis diagnosis based on ICD-8, 9 and 10.

| Characteristic | Full Sample<br>(N = 661,537) |  | Full Siblings<br>(N = 236,414) |  | Cousins<br>(N = 370,676) |  |
| --- | --- | --- | --- | --- | --- | --- |
|  | N | % | N | % | N | % |
| Postpartum psychosis diagnosis | 1,765 | 0.27 | 555 | 0.23 | 1,105 | 0.30 |
| Inpatient postpartum psychosis diagnosis <sup>a</sup> | 1,188 | 67 | 382 | 69 | 708 | 64 |
| Outpatient postpartum psychosis diagnosis <sup>a</sup> | 577 | 33 | 173 | 31 | 397 | 36 |

<sup>a</sup> Percentage breakdown of the diagnoses is the percentage out of the total number of postpartum psychosis cases.

As a next step, we limited the sample to individuals with ICD-10 diagnosis codes only and re-estimated the relative recurrence risk. We excluded all women who gave birth prior to January 1997 in our dataset, as ICD-10 was implemented in Sweden in 1997. This subcohort included 790,300 women, of whom 1,871 (0.24%) experienced postpartum psychosis (Table 3). We estimated a full sibling relative recurrence risk of 8.65 (95% CI=4.58–14.72; p<0.001; Tables 4 and S3). We could not estimate the risk among cousins since there were no concordant cousin pairs.

**Table 3.** Sample characteristics of mothers with a first-ever childbirth between January 1, 1997, and October 31, 2017, and postpartum psychosis diagnosis based on ICD-10.

| Characteristic | Full Sample<br>(N = 790,300) |  | Full Siblings<br>(N = 306,270) |  | Cousins<br>(N = 412,662) |  |
| --- | --- | --- | --- | --- | --- | --- |
|  | N | % | N | % | N | % |
| Postpartum psychosis diagnosis | 1,871 | 0.24 | 640 | 0.21 | 1,156 | 0.28 |
| Inpatient postpartum psychosis diagnosis <sup>a</sup> | 1,294 | 69 | 447 | 70 | 747 | 65 |
| Outpatient postpartum psychosis diagnosis <sup>a</sup> | 577 | 31 | 193 | 30 | 409 | 35 |

<sup>a</sup> Percentage breakdown of the diagnoses is the percentage out of the total number of postpartum psychosis cases.

**Table 4.** Relative recurrence risks for postpartum psychosis among female full siblings and cousins of mothers.

| Model and sample type/diagnosis type | Full Siblings |  |  | Cousins |  |  |
| --- | --- | --- | --- | --- | --- | --- |
|  | RRR | 95% CI | P | RRR | 95% CI | P |
| **Adjusted model for birthyear <sup>a</sup>**<br>Inpatient and outpatient | 10.34 | 6.58–16.20 | <0.001 | 1.78 | 0.70–3.62 | 0.161 |
| **Adjusted model for birthyear and age at childbirth**<br>Inpatient and outpatient | 10.69 | 6.60–16.26 | <0.001 | 1.78 | 0.70–3.62 | 0.162 |
| **Adjusted model for birthyear, age at birth, and history of bipolar disorder diagnosis in the sibling/cousin before first-ever childbirth**<br>Inpatient and outpatient | 6.88 | 3.50–12.71 | <0.001 | 1.44 | 0.51–3.54 | 0.453 |
| **Adjusted model for birthyear and age at childbirth, after 2001**<br>Inpatient and outpatient | 7.82 | 3.88–13.95 | <0.001 | 1.79 | 0.71–3.66 | 0.141 |
| **Adjusted model for birthyear and age at childbirth, after 2001**<br>Inpatient | 6.75 | 2.66–13.90 | <0.001 | 1.40 | 0.35–3.65 | 0.563 |
| **Adjusted model for birthyear and age at childbirth**<br>Inpatient and outpatient<br>ICD-10 diagnoses only | 8.65 | 4.58–14.72 | <0.001 | - | - | - |

<sup>a</sup> Birthyear, year of first-ever childbirth.
RRR, relative recurrence risk; 95% CI, 95% confidence intervals; ICD-10, International Classification of Diseases Tenth Revision.
Note: We could not estimate the risk among cousins since there were no concordant cousin pairs.

As demonstrated, the prevalence of postpartum psychosis was observed to be 0.15%, following the first childbirth and up to three months post-pregnancy. This rate effectively signifies the absolute risk of developing postpartum psychosis during this critical period among new mothers across the general population. Furthermore, with the relative recurrence risk of postpartum psychosis among female full siblings adjusted for birthyear identified as 10.34, the absolute risk for women with an affected sibling escalates to approximately 1.55% within the entire population. Therefore, despite the general risk of postpartum psychosis being relatively low at 0.15%, individuals with a familial history of the condition, particularly those with an affected sibling, experience a substantially elevated risk, though their absolute risk remains comparatively low, at an estimated 1.55% within the entire population.

## 4. Discussion

Using population-based Swedish register data, we estimated a prevalence of 0.15% for postpartum psychosis, which is comparable to previous Swedish register reports of 0.12% between 1975-2003 (23) and 1983-2000 (21).

To gain a deeper understanding of the risk architecture of postpartum psychosis, our study has conducted an analysis of familial risk. We found that female (i.e., female same-sex) full siblings of women experiencing their first episode of postpartum psychosis after pregnancy — termed ‘first-onset postpartum psychosis’ in our study — have an increased relative recurrence risk for the disorder. Full siblings, sharing approximately 50% of their segregating genes, also experience shared environments, environmental factors that siblings commonly experience, especially during their early developmental stages. Therefore, the increased disorder risk among full siblings can likely be attributed to a combination of both genetic factors and shared environment. Shared environment refers to environmental factors that siblings who live together commonly experience. Shared environment makes family members similar to one another beyond what can be explained by genetic influence alone.

The relative risk of postpartum psychosis for siblings was 10.69 (95% CI=6.60–16.26). In comparison, the familial recurrence risks documented for schizophrenia is 9.0 (95% CI=8.5-11.6) and bipolar disorder 7.9, (95% CI=7.1-8.8) in Swedish registry studies (24). This emphasizes the exceptional familial and potentially genetic predisposition towards postpartum psychosis. However, it is important to note that the familial recurrence risk in schizophrenia or bipolar disorder is observed in studies with both male and female participants, which may affect direct comparisons due to the sex-specific occurrence of postpartum psychosis.

To further dissect the risk factors for postpartum psychosis and determine whether they are purely genetic, purely environmental, or a combination of both, our study expanded its analysis to include cousins. We found a relative risk of 1.78, however, the confidence intervals for the relative recurrence risk in cousins were wide and included the value of one (95% CI=0.70–3.76). Therefore, the data thus did not allow us to separate the genetic effects from shared environmental influences.

We also demonstrated a similarly high full siblings relative recurrence risk when we restricted the analysis to ICD-10 codes to limit heterogeneity in diagnoses based on different ICD code definitions. In addition, the similarity in the risk estimates for female cousins between the overall and post-2001 analysis reassures us that our relative recurrence risk estimates accurately reflect the familial aggregation of postpartum psychosis. This indicates that any potential bias from missing data among cousins has been effectively addressed, allowing our findings to reliably represent the condition’s familial patterns without undue influence from the missing data.

We found that, in models adjusted for year of and age at childbirth and history of bipolar disorder before childbirth, the relative recurrence risk for female full siblings was statistically significant but lower than in models adjusted only for age at childbirth.

This reduction suggests the presence of confounding by the history of bipolar disorder. Confounding in this context means that the association between being a full sibling and the risk of postpartum psychosis is partly influenced by a shared predisposition to bipolar disorder, a known risk factor for postpartum psychosis. Importantly, the odds ratio of 2.08 08 (95% CI=1.03–4.01, p=0.036) for the history of bipolar disorder in our model reflects that having a female sibling with a history of bipolar disorder effectively doubles the risk of developing postpartum psychosis, underscoring both the direct influence of bipolar disorder as a risk factor and its indirect effects mediated through familial ties. This finding supports the idea that at least part of the increased risk for postpartum psychosis in full siblings can be attributed to shared factors, which likely include both genetic and environmental components, common to bipolar disorder and postpartum psychosis. This interpretation aligns with existing literature, which shows a high risk of postpartum relapse in women with a history of bipolar disorder (25) and with a recent study on polygenic risk scores in postpartum psychosis (13). In the study, elevated relative risk ratios for schizophrenia and bipolar disorder polygenic scores were observed in both women with first-onset postpartum psychosis and those with a history of bipolar disorder, compared to controls.

This study has multiple strengths. Our use of a population-based sample potentially limits selection bias. Previous studies of postpartum psychosis largely recruited small clinical samples of patients with postpartum psychosis and other pre-existing psychiatric diagnoses, which may result in patient self-selection bias.

However, register data also have several limitations. Firstly, as the data are available for diagnoses, there may be a time lag between the actual timing of disorder onset and patients seeking care. To include patients with a potential diagnostic delay in our sample, we selected a diagnostic period of 0-3 months postpartum for analyses. Secondly, the completeness of historical records presents a significant challenge, especially in identifying cousin relationships that necessitate data spanning multiple generations. This issue leads to instances of missing cousin relationships in our dataset. To mitigate the impact of varying data completeness across time, we have incorporated birthyear as a covariate in our analyses. Thirdly, our analysis, focused on families with female sibling pairs, may not fully represent the spectrum of familial risk for postpartum psychosis due to excluding families with only one female sibling.

This exclusion could skew the estimated relative recurrence risk, limiting the generalizability of our findings.

Future studies could address whether the risk of postpartum psychosis onset differs in families where live birth was not an outcome of the pregnancy (25). We limited our sample to live births for two reasons. First, the Medical Birth Register has a more complete coverage of live births than of stillbirths, miscarriages, or abortions (14, 18). Second, we limited the sample to live births to avoid potential confounding with genetic or psychological factors, such as stress. Such additional factors may drive psychiatric episodes after pregnancy termination or stillbirth, but life stressors have not generally been found to increase risk of postpartum psychosis (26, 27). Further research on these risk factors is crucial for improved patient risk screenings.

Our results provide guidance for clinicians working with pregnant women with personal or family histories of postpartum psychosis. Our findings suggest that female siblings of women who have experienced postpartum psychosis should be counselled that their own risk is elevated, but that the absolute risk for the disorder remains small. Furthermore, the finding of a potential genetic contribution to postpartum psychosis risk suggests that heritable factors should also be considered in future studies of causes and precipitants of postpartum psychotic episodes.

## Data Availability

Data may be obtained from a third party and are not publicly available. Data cannot be shared publicly owing to restrictions by law. Data are available from the National Medical Registries in Sweden after approval by the Swedish Ethical Review Authority.

## Author Contributions

Study concept and design: VB, APK, BM

Acquisition, analysis, or interpretation of data: VB, KH, APK, XL, BM, TMO, TKR

Drafting of the manuscript: VB, APK, BM

Critical revision of the manuscript for important intellectual content: All authors Statistical analysis: APK, BM

Obtained funding: VB, BM Study supervision: VB, BM

Data Sharing

## Acknowledgments

This study was supported by a grant from the Beatrice and Samuel A. Seaver Foundation (BM, APK); the National Institute of Mental Health (NIMH), R21MH131933 (VB, KH, APK, BM, TMO, TKR), R01 HD111117 (TKR); and the 2020 NARSAD Young Investigator Grant from the Brain & Behavior Research Foundation (BM, grant number 29355). The computation was performed on resources provided by SNIC through Uppsala Multidisciplinary Center for Advanced Computational Science (UPPMAX) under Project SENS-2018-588.

