## Supplemental Tables 1-4, Figures 1-2 for "Familial risk of postpartum psychosis"

### *Supplemental Information*

**Table S1.** International Statistical Classification of Diseases and Related Health Problems Eighth (ICD-8), Ninth (ICD-9), and Tenth Revision (ICD-10) codes for psychiatric diagnoses consistent with symptomology of postpartum psychosis, given to mothers 0-3 months following childbirth.

| Disorder name | ICD-8 code | ICD-9 Swedish version | ICD-10 code |
|---|---|---|---|
| Brief psychotic disorder | | | F23 |
| Psychotic disorders not due to a substance or known physiological conditions | 298.00–299.99 | 298A–X | F28, F29 |
| Manic episode | 296.1, 296.8, 298.1, 296.9 | 296A, 296G, 298B | F30 |
| Bipolar disorder | 296.1, 296.3, 296.8 | 296A/C/D/E/W | F31 |
| Major depressive disorder, single episode, severe with psychotic features | | | F32.3 |
| Recurrent depressive disorder, current episode severe with psychotic symptoms features | | | F33.3 |
| Puerperal psychosis | 294.4 | | F53.1 |

ICD-8: The eighth revision of the ICD was implemented in Sweden in 1968. ICD-9: The ninth revision of the ICD was implemented in Sweden in 1987. ICD-10: The tenth revision of the ICD was implemented in Sweden in 1997.
Swedish version refers to the national version of ICD codes used in Sweden and released by the Swedish National Board of Health and Welfare (Socialstyrelsen).

**Table S2.** International Statistical Classification of Diseases and Related Health Problems Eighth (ICD-8), Ninth (ICD-9), and Tenth Revision (ICD-10) codes for all psychiatric diagnoses

| **Diseases** | **ICD-8** | **ICD-9** | **ICD-10** |
|---|---|---|---|
| Any psychiatric disorder | 290–315 | 290–315 | F** (All codes starting with F) |

**Table S3.** Relative recurrence risks for postpartum psychosis among female full siblings and same-sex cousins of mothers with the diagnosis of postpartum psychosis.

| | **Unadjusted model Inpatient and outpatient** | | | **Adjusted model for birthyear Inpatient and outpatient** | | | **Adjusted model for birthyear and age at childbirth Inpatient and outpatient** | | | **Adjusted model for birthyear and history of BP Inpatient and outpatient** | | | **Adjusted model for birthyear and age at childbirth Inpatient and outpatient – ICD 10** | | |
|---|---|---|---|---|---|---|---|---|---|---|---|---|---|---|---|
| **Relatedness** Variable | Coefficient | RRR (95% CI) | P value | Coefficient | RRR (95% CI) | P value | Coefficient | RRR (95% CI) | P value | Coefficient | RRR (95% CI) | P value | Coefficient | RRR (95% CI) | P value |
| **Full siblings** | | | | | | | | | | | | | | | |
| Intercept | -6.65 | | <0.001 | -74.19 | | <0.001 | -74.70 | | <0.001 | -73.93 | | <0.001 | -52.56 | | <0.001 |
| Birthyear | - | - | - | 0.03 | 1.03 (1.03–1.04) | <0.001 | 0.03 | 1.04 (1.03–1.04) | <0.001 | 0.03 | 1.03 (1.03–1.04) | <0.001 | 0.02 | 1.02 (1.01–1.04) | <0.001 |
| Age at childbirth | - | - | - | - | - | - | 0.00 | 1.00 (0.99–1.01) | 0.409 | -0.00 | 1.00 (0.99–1.01) | 0.404 | 0.01 | 0.99 (0.97–1.00) | 0.061 |
| History of BP | - | - | - | - | - | - | - | - | - | 0.73 | 2.08 (1.03–4.01) | 0.036 | - | - | - |
| PP diagnosis | 2.61 | 13.59 (8.41-20.7) | <0.001 | 2.36 | 10.34 (6.58–16.20) | <0.001 | 2.37 | 10.69 (6.60–16.26) | <0.001 | 1.93 | 6.88 (3.50–12.71) | <0.001 | 2.16 | 8.65 (4.58–14.72) | <0.001 |
| **All cousins** | | | | | | | | | | | | | | | |
| Intercept | -5.92 | | <0.001 | -19.77 | | 0.031 | -20.17 | | 0.031 | -19.83 | | 0.034 | - | - | - |
| Birthyear | | | | 0.00 | 1.00 (1.00–1.02) | 0.131 | 0.00 | 1.00 (1.00–1.02) | 0.128 | 0.00 | 1.01 (1.00–1.02) | 0.137 | | | |
| Age at childbirth | | | | - | - | - | 0.00 | 1.00 (0.99–1.01) | 0.823 | 0.00 | 1.00 (0.99–1.01) | 0.829 | - | - | - |
| History of BP | | | | - | - | - | - | - | - | 0.30 | 1.34 (0.62–2.57) | 0.413 | - | - | - |
| PP diagnosis | 0.60 | 1.82 (0.72-3.72) | 0.143 | 0.58 | 1.78 (0.70–3.62) | 0.161 | 0.57 | 1.78 (0.70–3.62) | 0.162 | 0.37 | 1.44 (0.51–3.52) | 0.453 | - | - | - |

BP, bipolar disorder; PP, postpartum psychosis.
RRR, relative recurrence risk; 95% CI, 95% confidence intervals.
Note: Results for the model adjusted for age at birth in cousins with an ICD-10 diagnostic code only are not available due to no concordant cousin pairs available.

**Table S4.** Relative recurrence risks for postpartum psychosis among female full siblings and cousins, born after 2001.

| | **Adjusted model for birthyear Inpatient and outpatient After 2001** | | | **Adjusted model for birthyear and age at childbirth Inpatient After 2001** | | |
|---|---|---|---|---|---|---|
| **Relatedness** Variable | Coefficient | RRR (95% CI) | P value | Coefficient | RRR (95% CI) | P value |
| **Full siblings** | | | | | | |
| Intercept | -22.66 | | 0.229 | -24.26 | | 0.197 |
| Birthyear | 0.01 | 1.01 (0.99–1.03) | 0.369 | 0.01 | 1.01 (0.99–1.03) | 0.325 |
| Age at childbirth | -0.01 | 0.99 (0.97–1.01) | 0.221 | -0.01 | 0.99 (0.97–1.01) | 0.228 |
| Postpartum psychosis diagnosis | 2.06 | 7.82 (3.88–13.95) | <0.001 | 1.91 | 6.75 (2.66–13.90) | <0.001 |
| **All cousins** | | | | | | |
| Intercept | -21.86 | | 0.125 | -21.86 | | 0.117 |
| Birthyear | 0.01 | 1.01 (0.99–1.02) | 0.254 | 0.00 | 1.01 (0.99–1.02) | 0.241 |
| Age at childbirth | -0.01 | 0.99 (0.98–1.01) | 0.269 | -0.01 | 0.99 (0.98–1.01) | 0.266 |
| Postpartum psychosis diagnosis | 0.58 | 1.79 (0.71–3.66) | 0.155 | 0.58 | 1.40 (0.35–3.65) | 0.563 |

RRR, relative recurrence risk; 95% CI, 95% confidence intervals.

**Figure S1.** Birthyear for all female full siblings and all cousins.

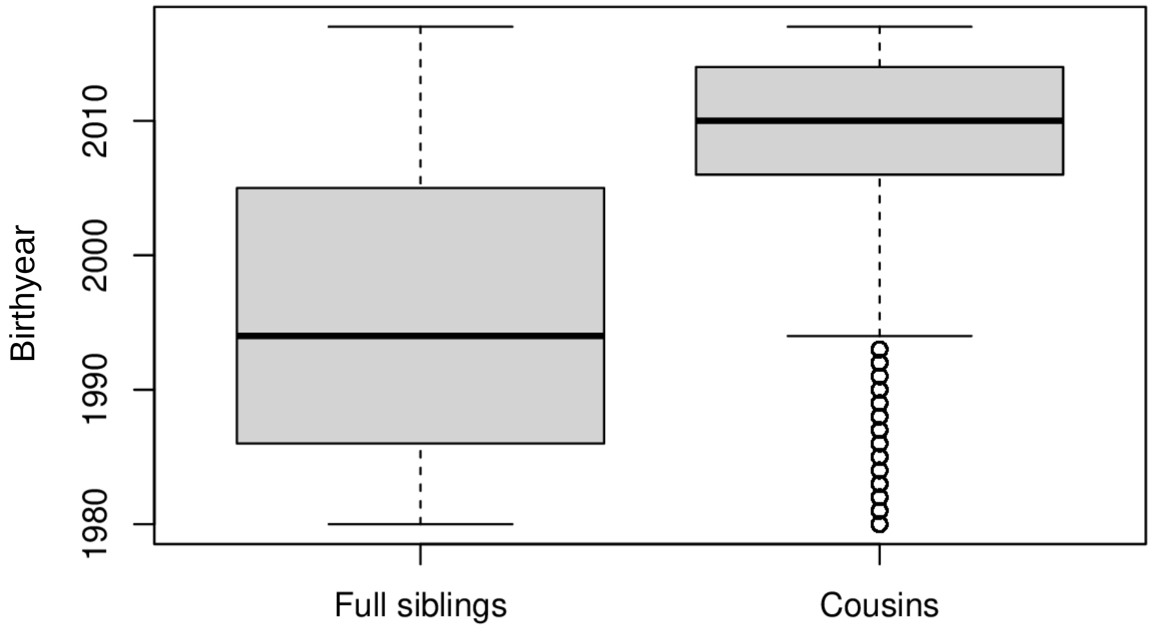

**Figure S2.** Postpartum psychosis prevalence estimates from 1980-2017, separated by overall, inpatient care, and outpatient care.

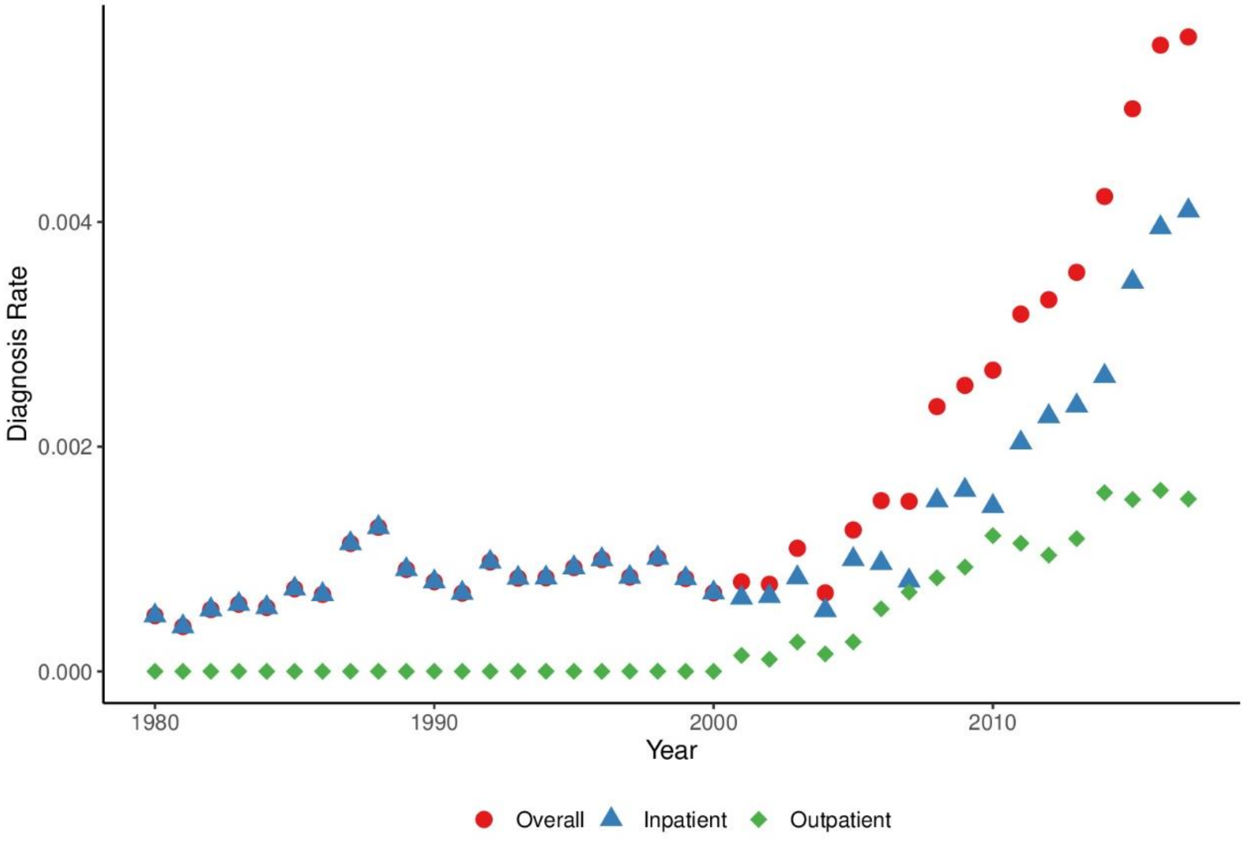


Note: Outpatient data available since 2001.
